# Automated human tracheal morphometrics using deep learning: Toward custom tracheostomy tubes

**DOI:** 10.1101/2025.11.27.25341143

**Authors:** Bram Ton, Sally Farooq, Jelmer Veenstra

## Abstract

**Purpose:** Tracheostomy is a critical, life-saving procedure, but complications often arise due to suboptimal tracheostomy tube fit. Customised tubes could mitigate these risks, yet their development hinges on precise tracheal morphometric data. This study aims to establish an automated workflow for extracting high-resolution tracheal measurements from CT scans to support the design of demographic-specific tracheostomy tubes.

**Methods:** This retrospective study utilised 100 anonymised CT scans, with 90 included after exclusions. An automated workflow was developed using 3D Slicer and a deep learning segmentation model (VISTA-3D) to extract tracheal measurements. Key parameters analysed included tracheal length, anteroposterior and transverse dimensions, and maximum inscribed circle diameter.

**Results:** Automated measurements revealed the following average tracheal dimensions:

- Length: 66.1 ± 12.2 mm
- Anteroposterior dimension: 16.2 ± 2.5 mm
- Transverse dimension: 17.9 ± 2.4 mm
- Maximum inscribed circle diameter: 14.1 ± 1.9 mm

Sex-specific differences were observed, with males exhibiting larger tracheal dimensions across all metrics.

**Conclusion:** The results align with existing literature, validating the automated approach for efficient, objective, and high-resolution morphometric analysis. This method supports the development of customised tracheostomy tubes tailored to demographic-specific needs, ultimately improving patient wellbeing.

## 1. Introduction

Tracheostomy is a surgical technique in which an opening is created in the anterior wall of the trachea to facilitate airflow, often in the context of upper airway obstruction. It can be performed either via an open surgical technique or percutaneous techniques. Indications can be emergent, such as in cases of acute upper airway obstruction or traumatic injury, or elective, for example in cases of chronic aspiration, neurological conditions, or prolonged ventilator dependence [1].

Reported rates of complications vary greatly [2]. Complications of tracheostomy are generally divided into transoperative, early, and late. Transoperative complications include haemorrhage (most common), pneumothorax, injury to the recurrent laryngeal nerve, oesophageal perforation [3] and intraoperative loss of airway [4]. Loss of airway can result from obstruction of the tracheostomy tube lumen by blood clots or mucus.

A number of the late complications of tracheostomy arise from prolonged contact between the cannula and the tracheal wall, resulting in local trauma. Continuous friction is thought to cause granulation tissue formation, which can mature to form a fibrotic tissue layer, potentially resulting in tracheal stenosis. Tracheal stenosis resulting in lumen occlusion from 60-70% to total occlusion is particularly unfortunate [4]. Reported rates of clinically significant tracheal stenosis range from 3 to 12% [5]. The use of an oversized tube is a risk factor [5]. Malposition of the distal cannula tip against the posterior tracheal wall is another risk factor for trauma, a problem especially relevant in obese patients with increased neck thickness [3]. Chronic irritation can also lead to chondritis, weakening the tracheal rings and potentially resulting in tracheomalacia [3]. Among the most severe late complications is tracheo-innominate fistula, a rare but life-threatening event. It develops from local trauma to the anterior tracheal wall [6] and excessive cannula movement [3] leading to erosion into the innominate artery [3, 6]. The risk is particularly high when the tracheostomy is placed low (below the 3rd third ring) [3]. Another uncommon late complication is tracheo-oesophageal fistula, which likely occurs secondary to chronic trauma and ischaemia of the posterior tracheal wall [3].

Choice of tracheostomy tube size is important to mitigate the risk of aforementioned complications. Parameters to consider include tracheal diameter, tracheal length (proximal, i.e. depth of soft tissue in the neck and distal), and the angle at which the tube sits in the trachea [7]. How well tube dimensions fit tracheal dimensions is therefore an important consideration, yet standard sizing assumes a relatively uniform anatomy despite the limited data available on normal adult tracheal measurements.

Ideally, each patient should have access to tailor-made tracheostomy tubes to ensure a perfect fit, and thus mitigate the risk of complications and increase the quality of life. Though many advances have been made in this direction, especially with the rise of additive manufacturing [8, 9], mass adoption is still far ahead. A compromise in this regard is to offer a set of products tailored to specific target groups.

To identify these target groups and obtain the optimal parameters, a large number of tracheal measurements have to be made. CT scans provide a valuable source of information to obtain these parameters. Obtaining representative measurements for each target group, requires large numbers of measurements. Obtaining these measurements manually would be too resource intensive, hence this article explores how these measurements can automated.

## 2. Materials and methods

### 2.1. Data

The initial data set consists of 100 CT scans provided by the Medisch Spectrum Twente (MST), The Netherlands. An internal ethics committee of the MST granted permission to use the data. A selection of recent adult neck scans was provided by the radiology department. The data was anonymised before being exported, only the year of birth and sex remained in the metadata. A demographic breakdown of the subjects included in this research is given in Figure 1. The figure shows that the data is skewed towards males and towards a slightly elderly part of the population.

**Figure 1.**
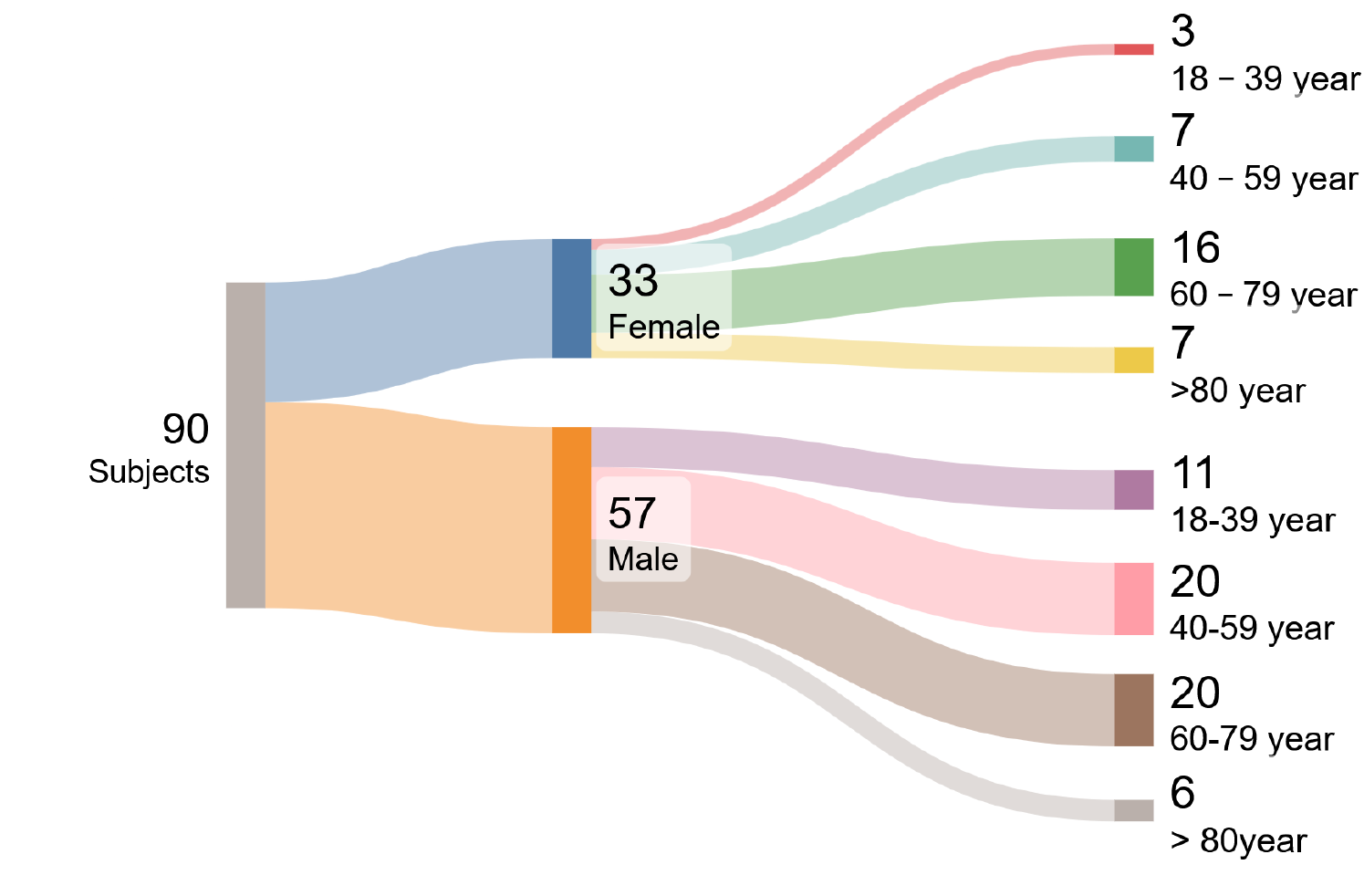
Demographic breakdown

### 2.2. Methods

The open source program Slicer was used [10] to automate the morphometric measurements of the trachea. This software was used because the processing steps available in the graphical user interface (GUI) can also be automated using Python code. The major advantage of this was that parameter settings and procedures could easily be evaluated visually in short iteration cycles. Thereafter these same steps can be added to the Python script to automate the workflow.

#### 2.2.1. Segmentation

The first step was the conversion of the data from the Digital Imaging and Communications in Medicine (DICOM) format to the Neuroimaging Informatics Technology Initiative (NIfTI) format. This was done using the `dcm2niix` tool. This conversion was required as the segmentation model works with this format.

For the segmentation itself, the VISTA-3D segmentation model [11] was used. This segmentation model is a deep learning based foundation model capable of segmenting 127 classes from CT scan data, including the trachea and thyroid. The segmented trachea starts roughly at the thyroid cartilage and includes part of both primary bronchi. The segmentation results are represented by an integer label per voxel.

##### Cleaning

The segmentation process is not perfect, hence some minor cleaning operations are required next. The cleaning operations make use of the segmentation toolbox [12] of Slicer. The regions at the extents of the trachea can contain small noisy blobs which are part of the segmentation. To remove these, we retain the largest blob only. In the case of the thyroid, the segmentation sometimes misses the region around the isthmus. This implies that the left and right lobe are not always connected, hence the strategy of retaining only the largest blob will not work. In this case the strategy is to remove all blobs below a certain volume threshold. The threshold parameter is defined in voxels. Based on the image spacing and a target threshold volume of 1 cm^3^, the target number of voxels for the threshold is determined.

The segmented trachea sometimes contains small holes. These holes interfere with the determination of the cross sectional area. A morphological closing operation is performed using a 4 mm kernel to ‘fill’ these holes.

#### 2.2.2. Centreline

After segmentation and cleaning, the next step is to extract the centreline of the trachea. This centreline is used to define at which positions the cross sectional areas of the trachea will be determined. The Vascular Modeling Toolkit (VMTK) extension of Slicer is used for determining the centreline and calculating the cross sectional areas. The VMTK was designed for analysing blood vessels [13], but also works well for other tubular structures such as the trachea. This toolkit is used to extract the centrelines of the trachea, and usually results in at least three lines. One for the main part of the trachea and two for the left and right parts of the primary bronchi. The longest line, representing the main part of the trachea, is selected for further processing.

To be able to derive some statistics of the cross sectional areas, it is important that they are all measured at the same location along the centreline. The measurements start at a point just above the carina and move upward towards the larynx. The extracted centreline is resampled so that the new control points have a fixed distance of 5 mm.

##### Centreline cut-off

The centreline extraction near the extents of the segmented trachea are not well defined. This is caused by an irregular segmentation at these areas. To compare individual trachea to each other, it is important to define a consistent end-point along the centreline. A landmark used to determine this end-point is the plane which is perpendicular to the centreline and just touches the lowest part of the segmented thyroid. The normal of this plane is defined by the vector starting at the carina (index 0) and pointing to the point which is 20 mm away from the carina (index 4). The point used in describing the plane is the coordinate of the lowest part of the segmented thyroid (Figure 2b).

**Figure 2.**
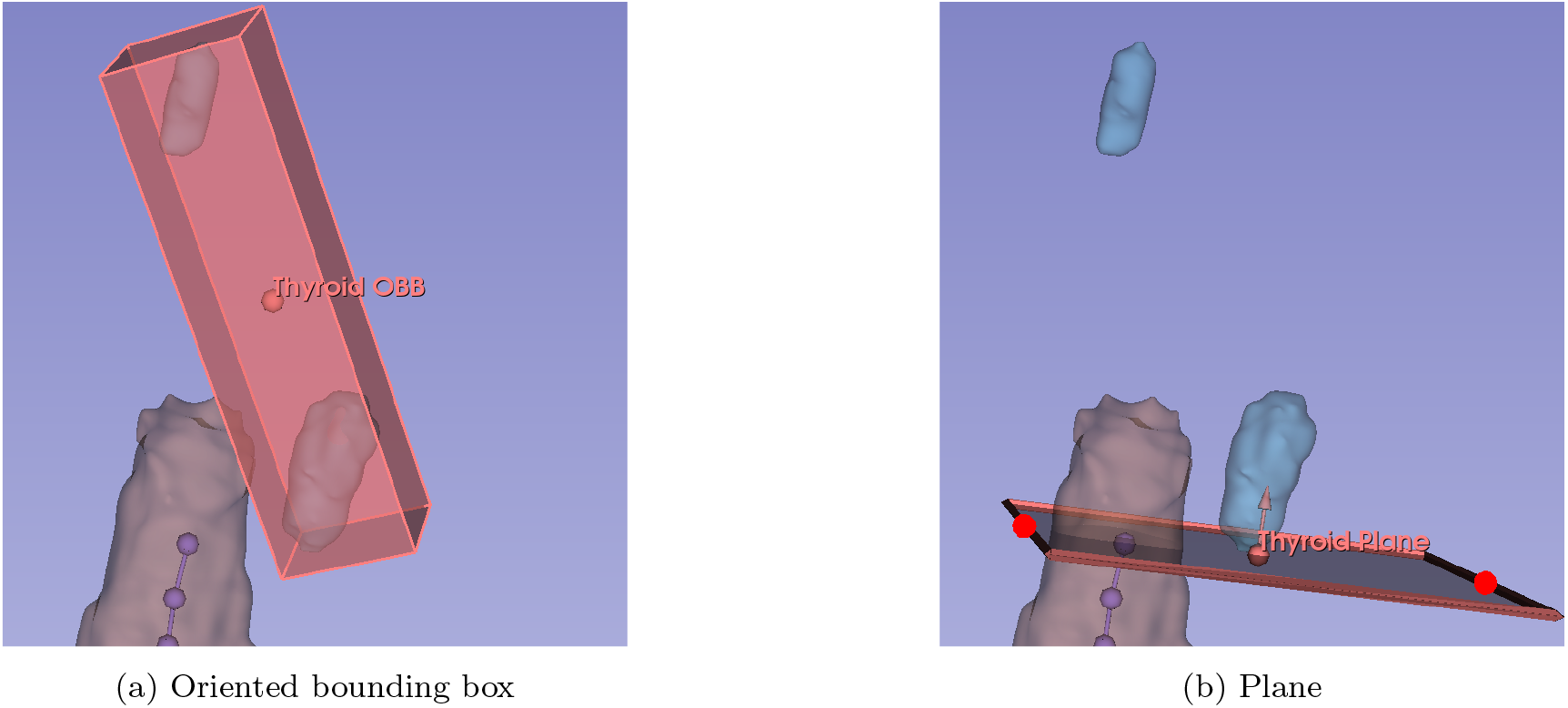
End-point landmark determination of tracheal centreline

Using this plane, the end-point is defined as the last point of the centreline which still resides below the defined plane.An initial approach to determine this end-point was to use the lowest plane of the oriented bounding box (Figure 2a) of the segmented thyroid. This approach was not successful as the segmentation of the thyroid was not always complete.

#### 2.2.3. Metrics

This section describes several of the metrics which are derived from the CT scans in an automated way. First is the cross-sectional area of the trachea at fixed intervals, thereafter the diameter of the maximum inscribed circle is described. At last the measurement of the anteroposterior and transverse dimensions are elaborated. Figure 3 provides an overview of how some of the metrics are determined. A cross section at each node of the centreline is defined, but in the figure only three are displayed as an example.

**Figure 3.**
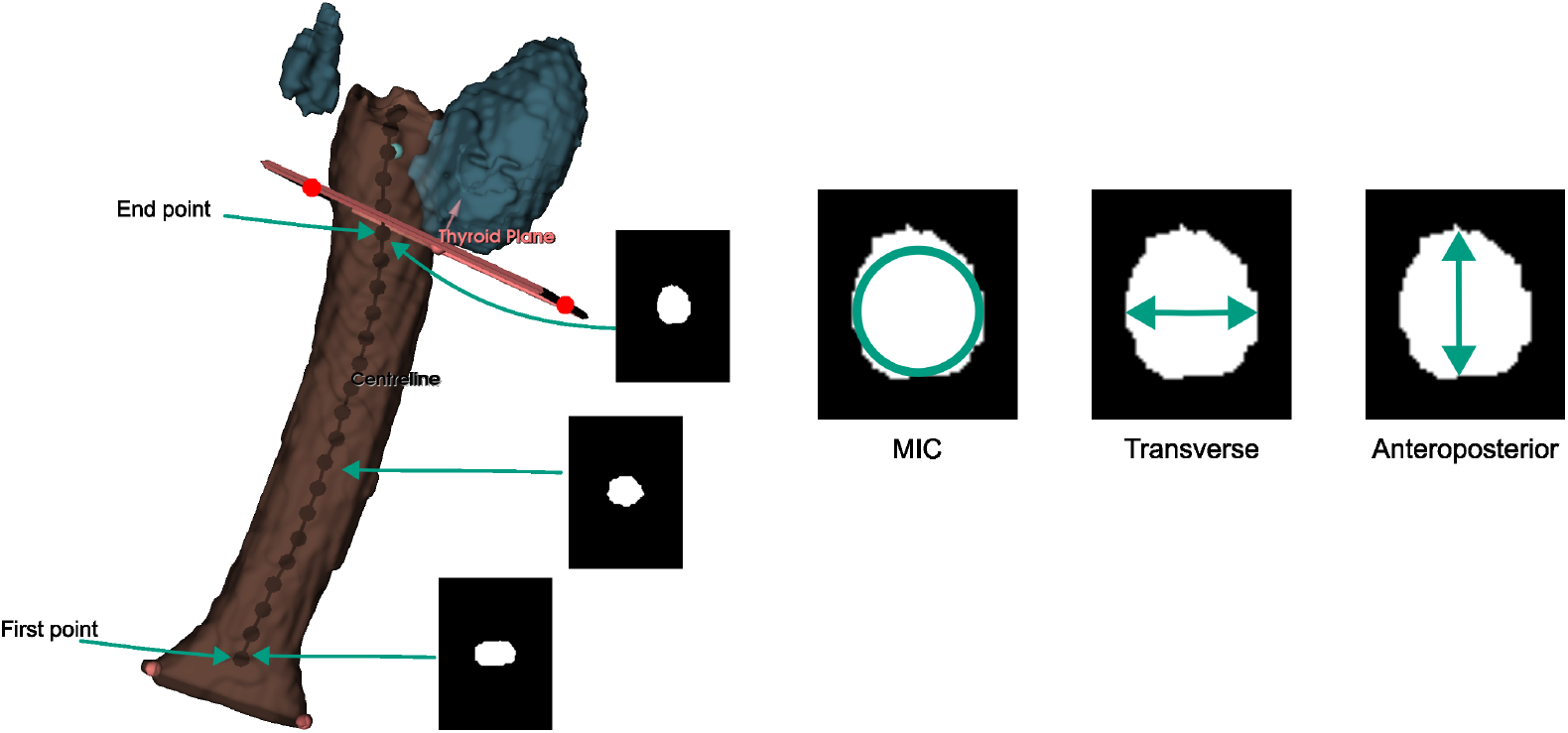
Centreline with examples of cross sections

##### Length

In our work we define the length of the trachea as the summation of all distances between control points of the centreline up to the previously defined end-point of the trachea (See Section 2.2.2).

##### Area

To obtain the actual cross sectional area, the internal workings of the toolkit define a cross cut perpendicular to the centreline at the position of the specified control point. The internal workings of the Slicer program can be called from within a Python script, hence it is possible to extract the cross sectional areas of the trachea in this way.

##### Maximum inscribed circle

The maximum inscribed circle (MIC) of is an important parameter as it directly relates to the diameter of the tracheostomy tube. The MIC is determined by first storing a binary image of the tracheal cross-section. Cross-sectional images are captured at each control point along the centreline of the trachea. After this a distance transform is applied to the binary images, the maximum value of this transform determines the radius of the maximum inscribed circle in pixels. The final step is a unit conversion from pixels to mm. This is done using the spacing information from the CT scan file.

##### Anteroposterior and Transverse dimensions

The same binary image can be used to determine the anteroposterior (AP) and the transverse (T) dimension of the trachea. To determine the AP dimension, the binary image is summed in the first dimension and the maximum value is used. To determine the transverse dimension, the binary image is summed in the second dimension and the maximum value is determine once more. Conversion from pixel units to mm is once again done using the spacing information from the CT file.

## 3. Results

### 3.1. Exclusion

Out of the 100 subjects analysed, it was not possible to define a conclusive centreline cut-off point for four of the subjects. Out of these, two subjects has an insufficient complete segmentation of the upper part of the trachea which made it impossible to find the last point of the centreline which still resided below the thyroid plane. In one case the subject was intubated during the CT scan, which led to a corrupted segmentation of the trachea. In the other case there was a paratracheal air cyst [14] present which confused the centreline finding algorithm. These four subjects are therefore not used for further analysis.

Each of the scans has also been manually inspected to verify if a significant part of the main bronchi was present in the scan. Presence of these bronchi is necessary to determine the point just above the carina. In four cases the bronchi was not present, and in two cases the bronchi part was not sufficient enough for the centreline algorithm to work. These six cases have also been excluded from further analysis. The total number of subjects included for statistical analysis is therefore now 90.

### 3.2. Metrics

This section dives into the details of the following metrics of the trachea: length, area, minimum inscribed circle (MIC), the anteroposterior dimension and the transverse dimension.

#### 3.2.1. Length

A histogram describing the trachea length is given in Figure 4. We see that the majority of the length measurements fall within the range 60–70 mm.

**Figure 4.**
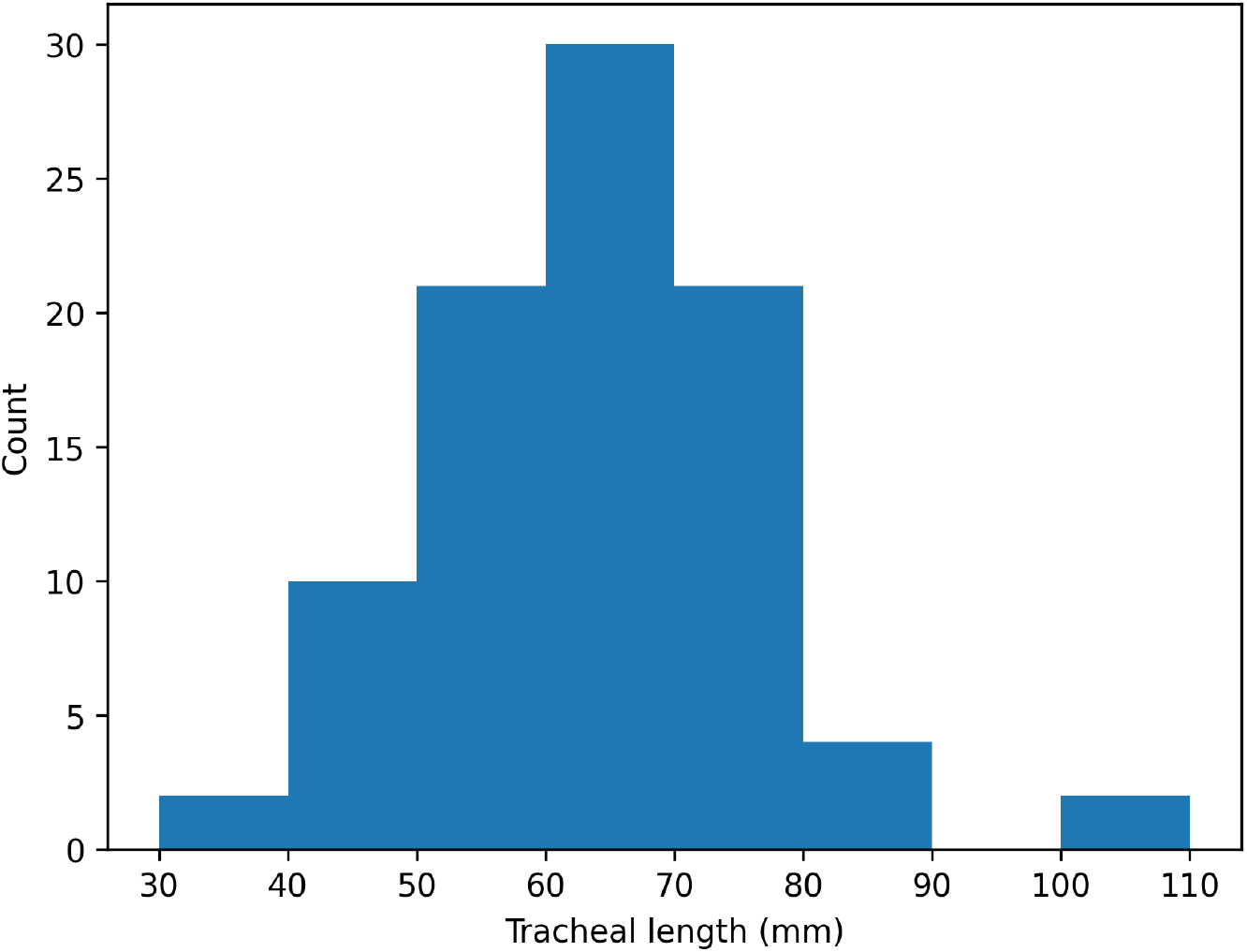
Tracheal length (mm)

#### 3.2.2. Area

The statistics of the cross-sectional area of the trachea can be seen in Figure 5. The figure shows a box plot per 5 mm along the centreline of the trachea. The number above the median line indicates the number of samples which were available for calculating the statistic.

**Figure 5.**
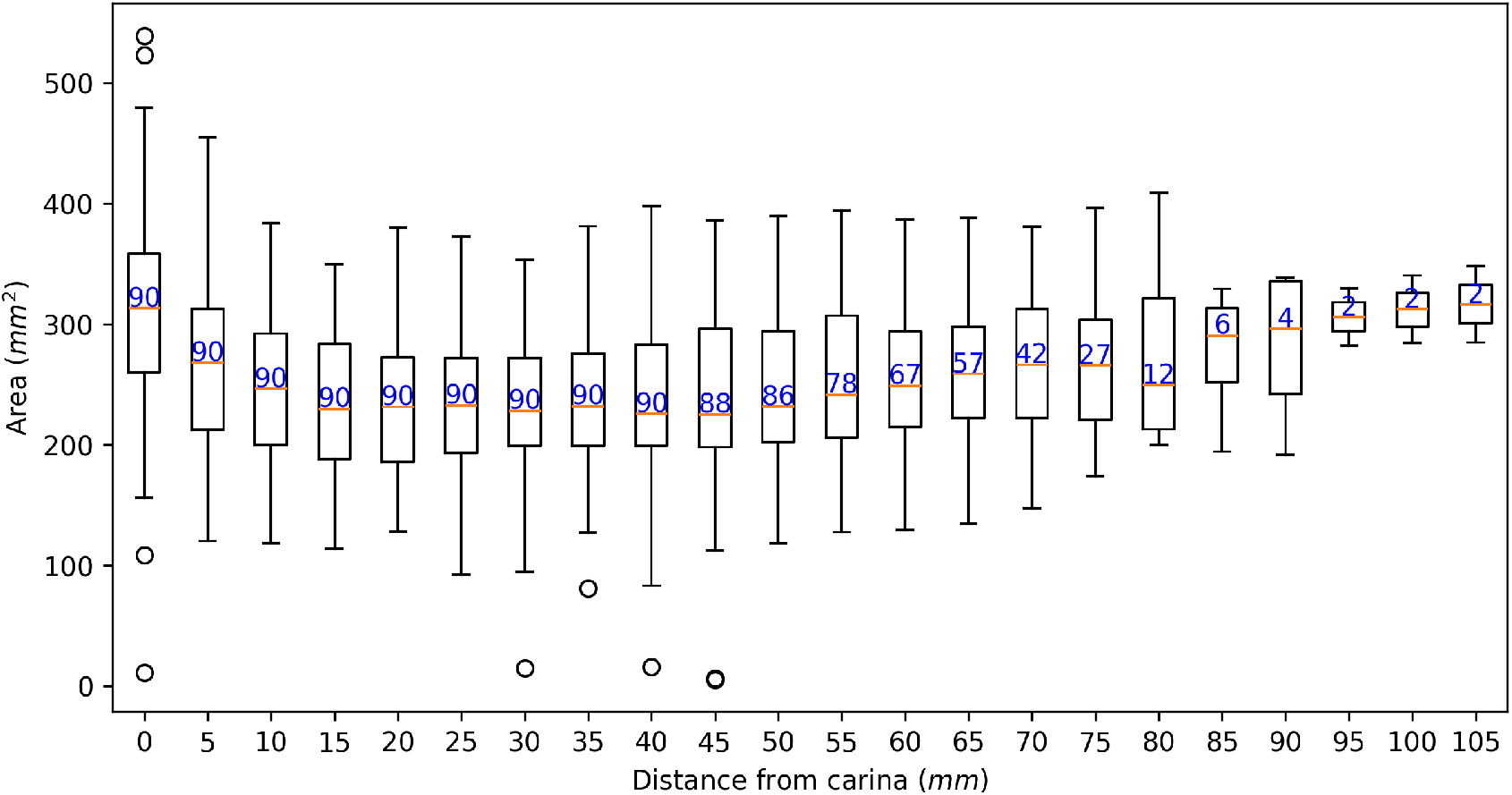
Cross-sectional area of the trachea (mm^2^)

The general trend of tracheal area which can be seen in the figure is a large area at the start, then drops, and then finally increases slowly again. The drop in the beginning can be explained by the fact that this is area corresponds to the region of bifurcation.

Similar plots could have been made from the other measurements, but have been omitted for conciseness. Main goal of this figure was to show that it is possible to obtain high spatial resolution along the trachea. The other metrics are summarised in the next section.

### 3.3. Summary

Table 1 summarises some of the automated morphometrics of the trachea. These values were computed by averaging the values along the trachea. The findings from the table confirm prior observations that females have a smaller trachea in general.

**Table 1:**
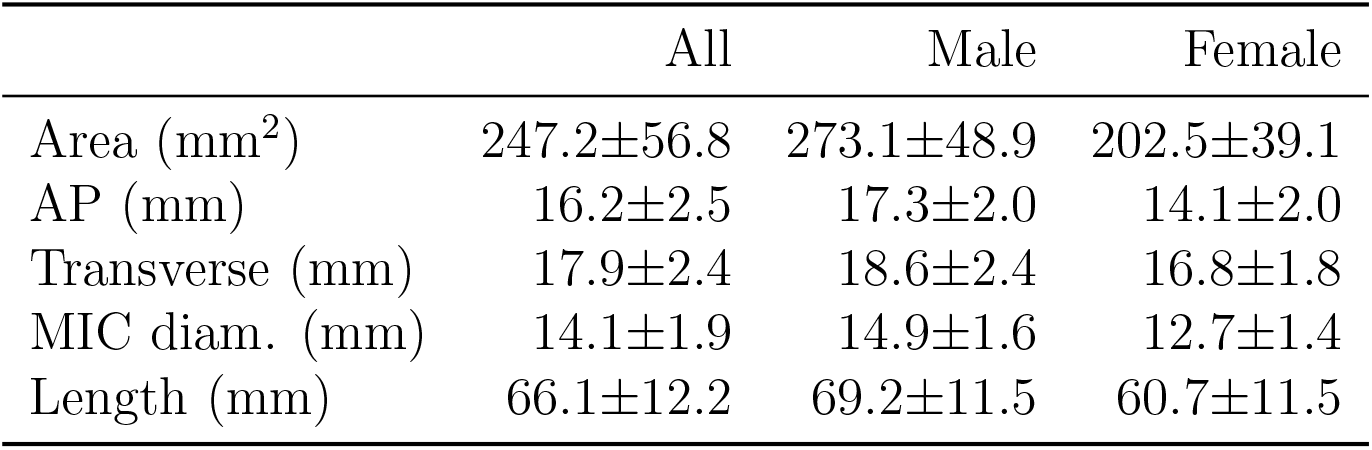
Morphometrics summarised.

The automated cross-sectional area reported by Ebrahimian et al. ranged from 204–273 mm^2^ for normal patients taking into account both pulmonary phases [15]. For this same group of the patients the AP dimension ranged from ≈ 16–18 mm. Our values show a good resemblance to these existing numbers.

If we look at the average male transverse dimension of the trachea we find good alignment with the value reported by Prince and Stark [16]. They report a dimension of 19.97 ± 2.37 mm.

The length of the trachea reported here is significantly shorter compared to values reported by Kamel et al., whom report an average tracheal length of 102.8 ± 9.9 mm. This large deviation can be explained by the fact that the work of Kamel uses the cricoid as a reference and we use the inferior of the thyroid as reference.

## 4. Discussion

A common landmark used with regards to tracheal measurements is the cricoid. In this work it was not possible to automatically determine this point hence the superior of the thyroid has been used. This affects the length measurement of the trachea.

This work used a foundational model to perform the segmentation. It is likely that better results can be obtained if this foundational model is fine tuned to increase its accuracy of the classes of interest such as the trachea and the thyroid. Furthermore, it is worthwhile to explore if the cricoid can be added as new class to the model. The cricoid is a common landmark on which trachea morphometrics are based.

Currently the normal of the cross sectional plane is determined with a simple heuristic based on the control points of the centreline. A more refined method would be to fit a curve through the control points, this would ensure a more smoothed line which can be used for determining cross sectional planes. Benefit of this approach would be that less accurate control points will not influence the determination of cross sectional planes too much.

### 4.1. Limitations

Our study has a few limitations. First limitation is that the set of CT scans used for analysis does not represent a valid cross section of the entire population. Only patients with a medical indication will be scanned. Furthermore, the sex distribution of the sample set used is skewed towards males. In our case the sample set consisted of 57 males and 33 females.

Another limitation is that a clear bifurcation of the trachea needs to be present in the data in order to obtain the reference point just above the carina.

An important factor for correct fitting of the tracheostomy tube is the skin to tracheal wall depth. This measurement is currently not available, but can easily be added.

## Data Availability

All data produced in the present study are available upon reasonable request to the authors.

## Declaration of generative AI and AI-assisted technologies in the manuscript preparation process

During the preparation of this work the author(s) used Mistral AI in order to draft an abstract. After using this tool/service, the author(s) reviewed and edited the content as needed and take(s) full responsibility for the content of the published article.

## Acknowledgements

We would like to thank the Slicer community, seldom have we seen such helpful (and civilised) responses on a discussion forum. We also thank the Medisch Spectrum Twente for providing the required data for our research and Atos Medical for their input.

## Research data

Due to the sensitive nature of the data used and the agreements made with the Medisch Spectrum Twente, sharing of data is not possible.

